# IL-6 signalling pathway inactivation with siltuximab in patients with COVID-19 respiratory failure: an observational cohort study

**DOI:** 10.1101/2020.04.01.20048561

**Authors:** Giuseppe Gritti, Federico Raimondi, Diego Ripamonti, Ivano Riva, Francesco Landi, Leonardo Alborghetti, Marco Frigeni, Marianna Damiani, Caterina Micò, Stefano Fagiuoli, Roberto Cosentini, Ferdinando Luca Lorini, Lucia Gandini, Luca Novelli, Jonathan P Morgan, Benjamin M J Owens, Karan J K Kanhai, Gordana Tonkovic Reljanovic, Marco Rizzi, Fabiano Di Marco, Alessandro Rambaldi

## Abstract

**Background:** Severe COVID-19 is characterised by interstitial pneumonia and hyperinflammation, with elevated levels of pro-inflammatory cytokines, such as IL-6. Effective treatments are urgently needed, and IL-6 is a rational target to reduce hyperinflammation.

**Methods:** An observational, control cohort, single-centre study initiated at the Papa Giovanni XXIII Hospital in Bergamo, Italy included patients with COVID-19 confirmed by a nasopharyngeal swab positive for severe acute respiratory syndrome coronavirus 2 RNA and interstitial pneumonia requiring ventilatory support. Patients were treated with either best supportive care and siltuximab or best supportive care alone. Propensity score matching was applied to minimise differences in baseline covariates between patient cohorts. The main outcome was mortality in siltuximab-treated patients compared with patients in the matched-control cohort. This study (<u>S</u>iltuximab <u>i</u>n <u>S</u>evere <u>CO</u>VID-19, SISCO) is registered with ClinicalTrials.gov, identifier NCT04322188.

**Findings:** Thirty patients received siltuximab, while 188 control patients received only best supportive care. Siltuximab-treated patients were matched to 30 control patients using the propensity score analysis of baseline covariates. The 30-day mortality rate was significantly lower in the siltuximab-treated than the matched-control cohort patients (HR 0·462, 95% CI 0·221– 0·965); p=0·0399). The mean follow-up was 33·3 days (range 7–58 days) for the siltuximab-treated patients and 22·8 days (range 2–45 days) for the control cohort. Sixteen siltuximab-treated patients were discharged from hospital, four remained on mechanical ventilation, and 10 patients died.

**Interpretation:** Patients with rapidly progressing COVID-19 respiratory failure requiring ventilatory support may benefit from treatment with siltuximab to reduce mortality and cytokine-driven hyperinflammation associated with severe disease. These findings require validation in a randomised controlled clinical trial.

**Funding:** Papa Giovanni XXIII Hospital and the Italian Association for Cancer Research funded the study. EUSA Pharma supplied siltuximab, and provided funding for data collection, analysis, and development of the publication.

## Introduction

Severe acute respiratory syndrome (SARS) and Middle East respiratory syndrome (MERS) are coronavirus infections characterised by rapid viral replication, massive inflammatory cell infiltration, and elevated levels of pro-inflammatory cytokines such as interleukin-1 (IL-1), IL-6, IL-8 and downstream proteins such as C-reactive protein (CRP), leading to acute respiratory distress syndrome (ARDS) and pulmonary injury.^1,2^ Similarly, infection with the severe acute respiratory syndrome coronavirus (SARS-CoV-2), the virus that causes corona virus disease, 2019 (COVID-19), also initiates hyperinflammation driven by elevated levels of cytokines.^3^ SARS-CoV-2 binds to alveolar epithelial cells, activating the immune system and resulting in an increase in the release of cytokines, including IL-6. These pro-inflammatory cytokines increase vascular permeability, leading to an increase in the amount of inflammatory exudates and blood cells entering the alveoli, resulting in dyspnoea and respiratory failure.^4^ Patients with severe or critical COVID-19 often develop severe interstitial pneumonia, and although the initial signs of inflammation are concentrated in the lungs, it can quickly progress to ARDS, systemic hyperinflammation, shock, and multi-organ failure leading to death.^5,6^ The systemic inflammation and hypoxic respiratory failure observed in severe or critical COVID-19 are associated with increases in IL-6 and CRP, which correlate with disease severity and patient prognosis.^7,8^ Elevated IL-6 levels have been shown to be predictive of respiratory failure, and elevated levels of both IL-6 and IL-8 have also been reported to be predictive of mortality.^8,9^

In addition to ventilatory support, targeting the IL-6 signalling pathway has been identified as a potential strategy to mitigate the elevated cytokines and resulting hyperinflammation associated with COVID-19.^6^ However, to date there are no data from randomised clinical trials and few controlled observational cohort studies describing the efficacy of therapies that target the IL-6 signalling pathway in patients with COVID-19.

Tocilizumab is an IL-6 receptor blocking monoclonal antibody (mAb) that is approved to treat elevated cytokines associated with the administration of chimaeric antigen receptor T-cell therapy, and received rapid approval in China for the treatment of patients with severe COVID-19 and extensive lung damage in March 2020.^10,11^ Reports from case series and observational studies suggest that tocilizumab has some activity in blocking the IL-6 receptor in patients with COVID-19, leading to improved oxygenation and a reduction in levels of CRP.^11-13^

In accordance with clinical guidelines developed at our medical centre, siltuximab was suggested as a treatment for hospitalised patients with COVID-19 and respiratory complications. Siltuximab is the first, and only FDA and EMA approved, mAb that specifically binds to IL-6 thereby inactivating IL-6–induced signalling. It is approved for the treatment of adults with multicentric Castleman disease who are human immunodeficiency virus and human herpes virus-8 negative.^14^ We examined the association between siltuximab treatment and mortality and/or respiratory function in patients hospitalised at the Papa Giovanni XXIII Hospital in Bergamo, Italy. We hypothesised that siltuximab would be associated with a low risk of mortality in analyses that were adjusted for major predictors of respiratory failure and weighted according to propensity scores to assess the drug’s efficacy.

## Methods

### Study design

On Mar 6, 2020, the Papa Giovanni XXIII Hospital in Bergamo, Italy made a request for siltuximab to be initially supplied under a compassionate-use programme for emergency treatment of patients with severe COVID-19. Consequently, an investigator-initiated study protocol was developed for immediate implementation. This prospective, observational cohort study was designed to report the outcomes of patients continuing to receive treatment with siltuximab under the compassionate-use programme, and retrospectively compare these outcomes to a matched-control group treated at the same centre. The study protocol was submitted and approved through the Hospital Ethics Board. Patients, or their legal representative, provided either verbal or written consent to participate in the study.

### Patients

Consecutive patients with COVID-19 and interstitial pneumonia, who were hospitalised and required ventilatory support by either invasive mechanic ventilation (IMV), non-invasive ventilation (NIV) or continuous positive airway pressure (CPAP) and who met the following criteria were included in the study: diagnosis of pulmonary infection with SARS-CoV-2 confirmed by a reverse-transcriptase quantitative PCR assay; and ARDS clinical picture in accordance with the Berlin 2012 criteria.^15^

Patients with an active bacterial or viral (not SARS-CoV-2) pulmonary infection not controlled by treatment were excluded from the study, and those treated with other anti-interleukin therapies, including IL-6 receptor blockers, were not eligible to participate. Those patients who were treated with siltuximab were grouped according to the type of ventilatory support received at the start of siltuximab treatment: patients receiving CPAP/NIV, and patients receiving IMV. The outcomes of patients treated with siltuximab were compared with those of patients from a control cohort who received best supportive care alone.

Patients in the control cohort were matched to the siltuximab-treated patients using a propensity score analysis. The control cohort patients were consecutively enrolled in the retrospective observational study (ReCOVID-19-2020) with the objective to describe and better understand the clinical profile of patients admitted to our institution. The ReCOVID-19-2020 study was approved by the Hospital Ethics board to collect demographic data, patient history, symptoms, laboratory abnormalities, radiological findings and clinical outcome data.

All patients were monitored according to the hospital and Italian national guidelines for a minimum of 30 days, and if a patient was discharged from hospital they were asked to provide relevant laboratory results and safety information for 30 days following the start of treatment.

### Study endpoints

The primary endpoint of this study was the reduction of mortality in COVID-19 patients treated with siltuximab compared with the matched-control cohort, calculated as the time from ventilation to death of any cause.

The secondary endpoints included the time to IMV or death in siltuximab-treated patients receiving CPAP/NIV at the time of treatment compared with the matched-control cohort. In addition, the ventilatory support parameters, respiratory function (ratio of arterial oxygen partial pressure to fractional inspired oxygen [PaO2/FiO2]), adverse events according to the National Cancer Institute Common Terminology Criteria for Adverse Events v4.03 within 30 days of treatment, and change in serum CRP levels (mg/dL) were described in all patients who received siltuximab. Any association or correlation between serum IL-6 (pg/mL) and mortality or prognosis was explored. The study protocol is available in the supplementary materials.

### Clinical and laboratory evaluation

Data were obtained from hospital medical records, and included demographic data, presenting symptoms and history of previous treatments, vital signs, ventilatory support, and laboratory data, including blood counts and aspartate aminotransferase (AST), alanine aminotransferase (ALT), creatinine, procalcitonin, lactate dehydrogenase, and IL-6, levels. Response to treatment was defined as the normalisation of CRP and blood counts, reduction in the need for ventilatory support, and the resolution of symptoms and signs of COVID-19.

### Confirmation of SARS-CoV-2 infection

Patients had nasopharyngeal swab sampling, testing for SARS-CoV-2 RNA using reverse-transcriptase quantitative PCR assay; patients with a positive test were considered confirmed cases of SARS-CoV-2 infection.

### Treatments

Patients treated with siltuximab received a dose of 11 mg/kg administered intravenously over an hour. A second dose was permitted 72 hours after the first dose at the physician’s discretion.

Best supportive care was provided according to hospital guidelines and included antiviral therapy (administration was influenced by drug availability and included lopinavir/ritonavir 200/50 mg two tablets twice daily, darunavir/cobicistat 800/150 mg one tablet once daily, or remdesivir 200 mg administered intravenously on day 1, followed by 100 mg daily for the remaining 9 days of treatment) and hydroxychloroquine 200 mg twice daily. Steroid use was not permitted according to local guidelines until March 27, 2020 when high-dose corticosteroids (intravenous methylprednisolone 1 mg/kg/daily for five days or equivalent dose of oral prednisone, both followed by tapering doses) were added to the treatment guidelines. Similarly, after that date, subcutaneous prophylactic low molecular weight heparin (4000 IU once daily) was introduced in all patients at admission.

### Statistical analyses

Logistic regression analyses using nearest neighbour propensity score matching were used to minimise differences in baseline covariates between patients in the control and siltuximab-treated cohorts. Patients in the control cohort were matched on a one-to-one basis with patients who received siltuximab using a number of baseline covariates at the start of ventilatory support, excluding treatment arm.

A two-sided Cox regression analysis was completed on the matched cohort analyses. For missing data points, the last observations were carried forward. All statistical analyses were performed with SAS^®^ software (Version 9.4 or later). Descriptive statistics are given separately for the two cohorts (CPAP/NIV and IMV) and for the whole population. The safety and endpoint analyses were based on the full-analysis set.

## Results

Thirty patients with COVID-19 confirmed by a nasopharyngeal swab positive for SARS-CoV-2 RNA and respiratory failure requiring ventilatory support were treated with siltuximab between March 7 and April 9, 2020, and 188 patients received best supportive care in the control cohort (ie, the ReCOVID-19-2020 study) between February 23 and March 13, 2020. Table S1 reports the baseline characteristics of all patients in the siltuximab-treated and control cohorts on the first day of ventilation.

### Propensity score analysis

We completed a nearest neighbour propensity score matching analysis. The following baseline covariates were identified in the model to select a total of 30 matches between the siltuximab-treated (n=30) and control cohort (n=30) patients: age, time from admission to initiation of ventilatory support, CPAP/NIV or IMV ventilation, logarithmic PaO2/FiO2, body mass index, comorbidities (hypertension, cardiovascular disease, cerebrovascular disease), and concomitant medication (antiplatelet therapy and angiotensin-converting enzyme [ACE] inhibitors). The standardised mean difference of the matched logit propensity scores for the baseline covariate was 0·0286, which is below the recommended calliper of 0·25 (figure S1). Tables 1 and 2 show that baseline demographic and laboratory parameters were similar between the siltuximab-treated and matched-control patients.

**Table 1:** Baseline patient and disease characteristics for the matched-control cohort and siltuximab-treated patients.

|  | **Siltuximab-treated patients, CPAP/NIV**<br>**n=25** | **Siltuximab-treated patients, IMV**<br>**n=5** | **All siltuximab patients**<br>**n=30** | **Control**<br>**n=30** |
| --- | --- | --- | --- | --- |
| **Sex** |  |  |  |  |
| Male | 19 (76.0%) | 4 (80.0%) | 23 (76.6%) | 24 (80.0%) |
| Female | 6 (24.0%) | 1 (20.0%) | 7 (23.3%) | 6 (20.0%) |
| **Age, years** |  |  |  |  |
| Median (IQR) | 64 (58–69) | 64 (53–65) | 64 (57–66) | 64.5 (56–70) |
| **Height, cm** |  |  |  |  |
| Median (IQR) | 175 (168–180) | 175 (170–180) | 175 (168–180) | 175 (172–180) |
| **Weight, kg** |  |  |  |  |
| Median (IQR) | 83 (77–91) | 87 (70–110) | 84 (77–95) | 83.5 (75–97) |
| **Comorbidities** |  |  |  |  |
| Hypertension | 11 (44.0%) | 1 (20.0%) | 12 (40.0%) | 11 (36.7%) |
| Diabetes | 6 (24.0%) | 0 | 6 (20.0%) | 5 (16.7%) |
| Cardiovascular disease | 4 (16.0%) | 0 | 4 (13.3%) | 4 (13.3%) |
| Malignancies | 2 (8.0%) | 1 (20.0%) | 3 (10.0%) | 0 |
| Cerebrovascular disease | 1 (4.0%) | 0 | 1 (3.3%) | 3 (10.0%) |
| Chronic kidney disease | 1 (4.0%) | 0 | 1 (3.3%) | 1 (3.3%) |
| **Medications at baseline** |  |  |  |  |
| Antiplatelet therapy | 4 (16.0%) | 0 | 4 (13.3%) | 6 (20.0%) |
| ACE inhibitors | 2 (8.0%) | 0 | 2 (6.7%) | 3 (10.0%) |
| Angiotensin receptor blocker | 8 (32.0%) | 1 (20.0%) | 9 (30.0%) | 5 (6.7%) |
| Anti-hypertensive therapy | 8 (32.0%) | 1 (20.0%) | 9 (30.0%) | 8 (26.7%) |
| Inhaler | 0 | 0 | 0 | 2 (6.7%) |
| Insulin | 1 (4.0%) | 0 | 1 (3.3%) | 2 (6.7%) |
| Oral anticoagulants | 0 | 0 | 0 | 2 (6.7%) |
| Oral antidiabetics | 5 (20.0%) | 0 | 5 (16.7%) | 3 (10.0%) |
| Proton pump inhibitors | 0 | 0 | 0 | 6 (20.0%) |
| Steroids | 0 | 0 | 0 | 2 (6.7%) |
| **Signs and symptoms** |  |  |  |  |
| Fever | 22 (88.0%) | 4 (80.0%) | 26 (86.7%) | 28 (93.3%) |
| Dry cough | 14 (56.0%) | 2 (40.0%) | 16 (53.3%) | 12 (40.0%) |
| Diarrhoea | 5 (20.0%) | 0 | 5 (16.7%) | 0 |
| Fatigue | 5 (20.0%) | 2 (40.0%) | 7 (23.3%) | 7 (23.3%) |
| Myalgia | 4 (16.0%) | 0 | 4 (13.3%) | 0 |
| Anorexia | 2 (8.0%) | 0 | 2 (6.7%) | 1 (3.3%) |
| **Respiratory support upon enrolment** |  |  |  |  |
| Invasive ventilation | 0 | 1 (20.0%) | 1 (3.3%) | 0 |
| Continuous positive airway pressure/non-invasive ventilation | 25 (100.0%) | 4 (80.0%) | 29 (96.7%) | 30 (100%) |
| **Time to start of ventilation from hospitalisation, days** |  |  |  |  |
| Median (IQR) | 2 (1–3) | 3 (3–3) | 2 (1–3) | 1 (1–2) |
| **PP arterial O<sub>2</sub>/fraction inspired O<sub>2</sub>** |  |  |  |  |
| Median (IQR) | 132.00<br>(92.86–154.00) | 96.00<br>(95.00–96.00) | 109.17<br>(92.86–153.00) | 121.50<br>(86.67–224.00) |

ACE=angiotensin-converting enzyme. IQR=interquartile range.

**Table 2:** Baseline laboratory and haematology parameters for the matched-control cohort and siltuximab-treated patients.

|  | **Siltuximab-treated patients, IMV**<br>**n=5** | **Siltuximab-treated patients, CPAP/NIV**<br>**n=25** | **All siltuximab patients**<br>**n=30** | **Control overall**<br>**n=30** |
| --- | --- | --- | --- | --- |
| **Haematological parameters** |  |  |  |  |
| **Median (IQR)** |  |  |  |  |
| WBC, median absolute number/μL (IQR) | 7450 (5500–16 870) | 8550 (6700–10 430) | 8515 (6700–10 470) | 8295 (6070–10 930) |
| Haemoglobin, median g/dL (IQR) | 14.70 (13.40–16.00) | 13.30 (12.60–14.30) | 13.40 (12.60–14.40) | 13.60 (11.50–14.80) |
| Platelets, median absolute number/μL (IQR) | 145 000 (140 000–153 000) | 223 000 (167 000–297 000) | 215 000 (147 000–279 000) | 189 000 (161 000–225 000) |
|  | **Siltuximab-treated patients, IMV**<br>**n=3** | **Siltuximab-treated patients, CPAP/NIV**<br>**n=22** | **All siltuximab patients**<br>**n=25** | **Control overall**<br>**n=27** |
| Lymphocytes, median absolute number/μL (IQR) | 860.0 (720.0–13 830.0) | 750.0 (500.0–880.0) | 760.0 (540.0–880.0) | 750.5 (552.5–1167.5) |
| Monocytes, median absolute number/μL (IQR) | 430.0 (360.0–610.0) | 285.0 (190.0–390.0) | 310.0 (200.0–390.0) | 427.7 (317.1–499.8) |
|  | **Siltuximab-treated patients, IMV**<br>**n=5** | **Siltuximab-treated patients, CPAP/NIV**<br>**n=23** | **All siltuximab patients**<br>**n=28** | **Control overall**<br>**n=28** |
| Neutrophils, median absolute number/μL (IQR) | 6140.0 (4180.0–12□500.0) | 7240.0 (5520.0–9140.0) | 7020.0 (5505.0–9320.0) | 7459.1 (4790.8–9198.0) |
| **Laboratory parameters median (IQR)** | **Siltuximab-treated patients, IMV n=5** | **Siltuximab-treated patients, CPAP/NIV n=25** | **All siltuximab patients n=30** | **Control overall n=30** |
| AST, median U/L (IQR) | 114 (47–114) | 56 (43–77) | 56 (43–89) | 62 (40–74) |
| ALT, median U/L (IQR) | 62.0 (51.0–107.0) | 56.0 (33.0–77.0) | 57.5 (33.0–92.0) | 36.0 (26.0–60.0) |
| Creatinine, median mg/dL (IQR) | 0.960 (0.740–1.010) | 0.850 (0.710–1.120) | 0.875 (0.710–1.090) | 0.955 (0.670–1.330) |
|  | **Siltuximab-treated patients, IMV n=2** | **Siltuximab-treated patients, CPAP/NIV n=6** | **All siltuximab patients n=8** | **Control overall n=14** |
| Procalcitonin, median ng/mL (IQR) | 0.445 (0.390–0.500) | 0.840 (0.200–3.460) | 0.445 (0.285–2.385) | 0.840 (0.470–3.080) |
|  | **Siltuximab-treated patients, IMV n=5** | **Siltuximab-treated patients, CPAP/NIV n=23** | **All siltuximab patients n=28** | **Control overall n=27** |
| Lactate dehydrogenase, median U/L (IQR) | 520.0 (390.0–579.0) | 500.0 (382.0–565.0) | 505.5 (384.0–567.0) | 503.0 (389.0–633.0) |
|  | **Siltuximab-treated patients, IMV n=4** | **Siltuximab-treated patients, CPAP/NIV n=24** | **All siltuximab patients n=28** |  |
| Interleukin-6, median pg/mL (IQR) | 446.07 (69.43–1842.63) | 118.95 (74.56–223.52) | 129.86 (74.56–237.88) |  |

|  | **Siltuximab-treated**<br>**patients, IMV**<br>**n=5** | **Siltuximab-treated**<br>**patients, CPAP/NIV**<br>**n=24** | **All siltuximab patients**<br>**n=29** | **Control overall**<br>**n=27** |
| --- | --- | --- | --- | --- |
| CRP, median mg/dL (IQR) | 9.90 (9.10–15.10) | 23.05 (16.35–25.80) | 19.90 (15.10–25.00) | 16.10 (12.70–28.70) |

alanine aminotransferase. AST=aspartate aminotransferase. CRP=C-reactive protein. IQR=interquartile range. WBC=white blood cells.
AL T=

### Siltuximab treatment

The majority of siltuximab-treated patients were receiving CPAP/NIV at the time of treatment. They were given a single dose of siltuximab within 48 hours of initiating ventilatory support (within 24 hours: 10/25 [40·0%]; within 48 hours: 19/25 [76·0%]). Five patients required IMV before siltuximab treatment. Of these patients, four progressed from CPAP/NIV to IMV before siltuximab treatment, and one required IMV upon admission and was then treated with siltuximab. Six patients received a second dose 72 hours after the first dose of siltuximab. Table S2 summarises doses of siltuximab administered during the study.

### Comparative endpoints

A total of 60 patients were included in the analyses for mortality, 30 in the siltuximab-treatment cohort and 30 in the matched-control cohort. The 30-day mortality rate was lower in patients treated with siltuximab than in the matched-control cohort (hazard ratio 0·462, 95% confidence interval [CI] 0·221–0·965); p=0·0399) (figure 1). The mean follow-up was 33·3 days (range 7–58 days) for the siltuximab-treated patients and 22·8 days (range 2–45 days) for the control cohort. Although not significant there was a trend toward a reduced need for mechanical ventilation in the 25 siltuximab-treated patients on CPAP/NIV compared with the 30 matched-control patients also on CPAP/NIV (figure 2; HR 0·615; 95% CI 0·362– 1·044).

**Figure 1:**
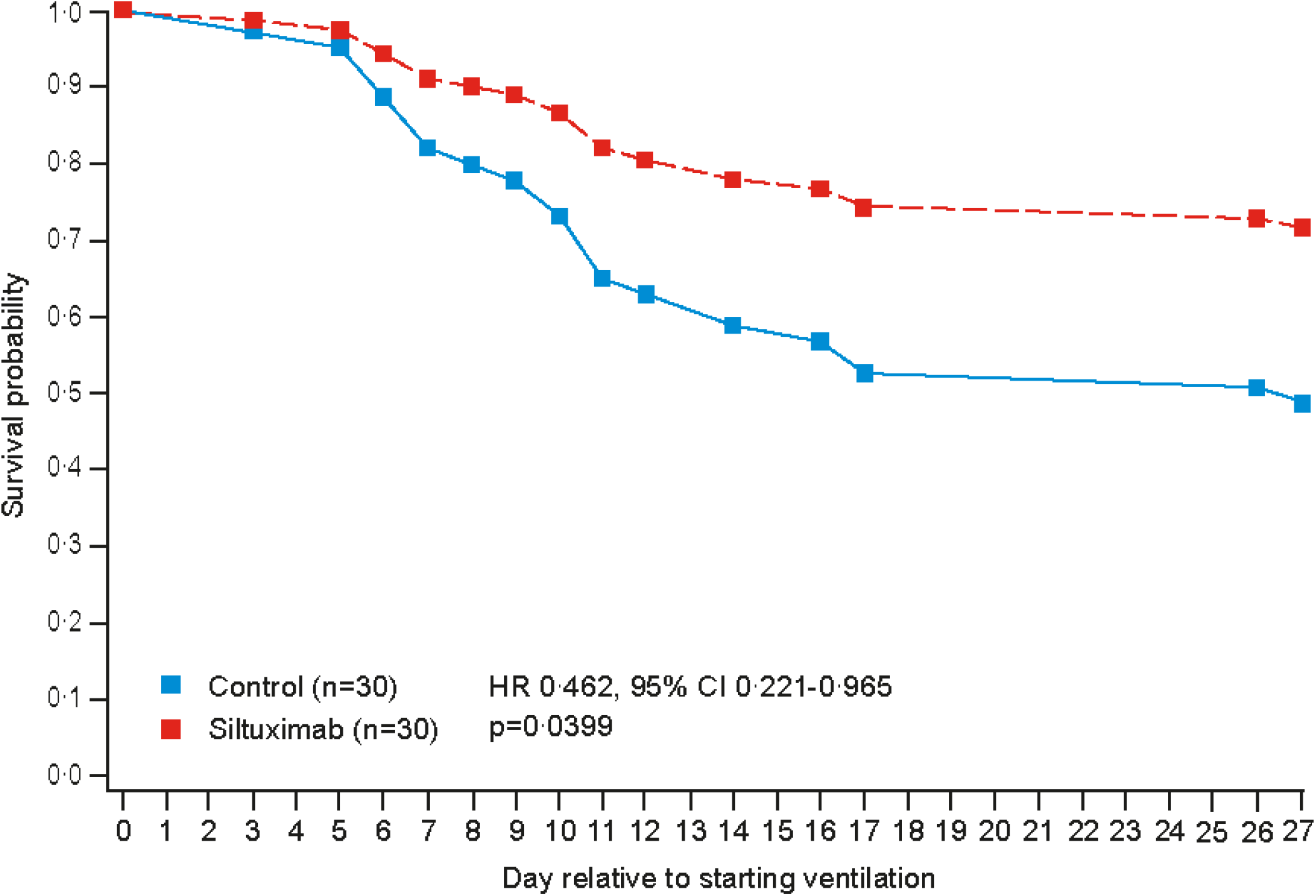
Probability of survival from the initiation of ventilatory support to death from any cause in siltuximab-treated patients (n=30) compared with control cohort (n=30)

**Figure 2:**
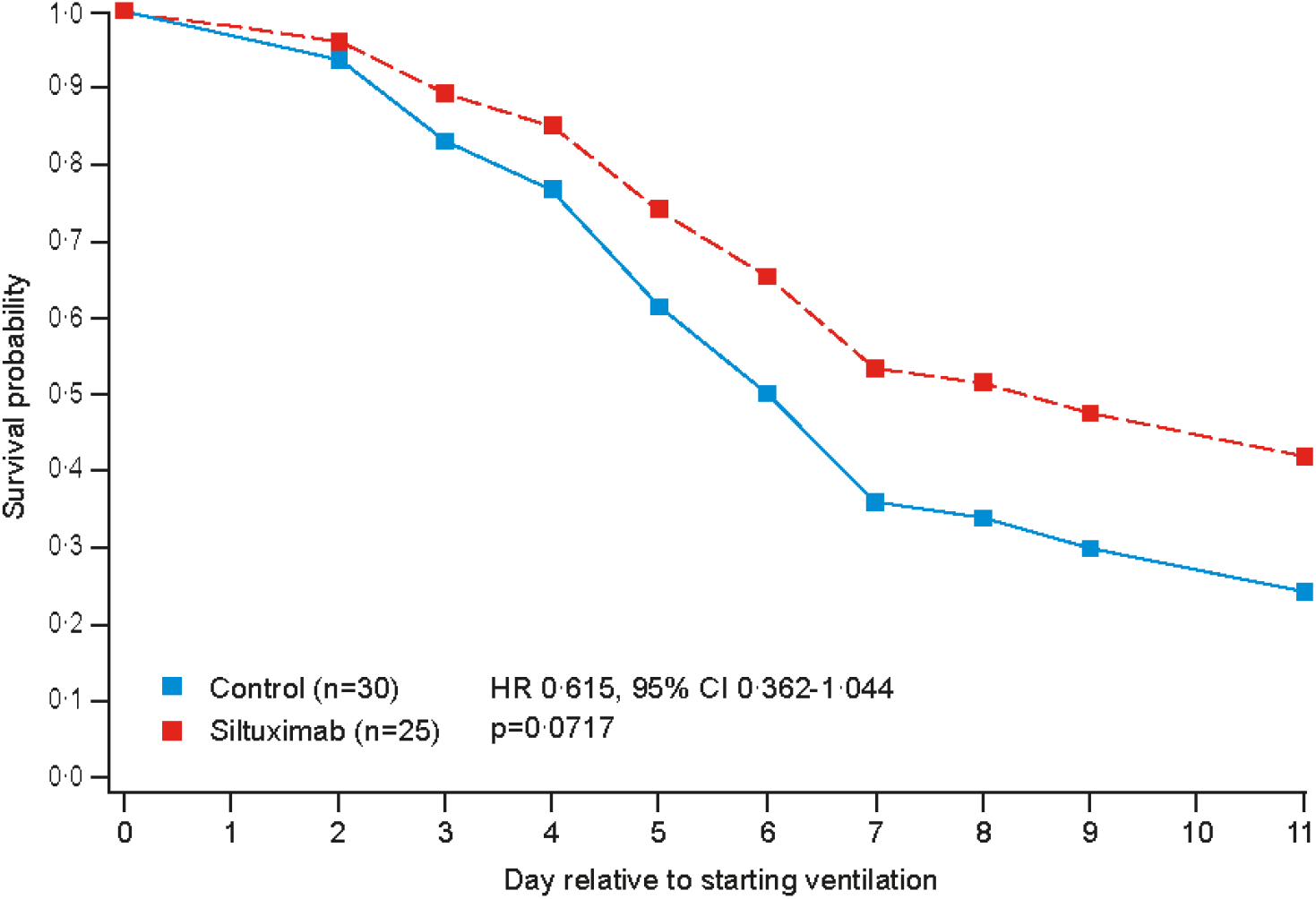
Probability of requiring mechanical ventilation or death in siltuximab-treated patients requiring CPAP/NIV (n=25) compared with the matched-control cohort (n=30)

### Non-comparative endpoints

Figure 3 shows changes in oxygen support and outcomes for all patients treated with siltuximab. At study end, of the 30 patients 16 (53·3%) recovered and were discharged, and four patients (13·3%) were on IMV.

**Figure 3:**
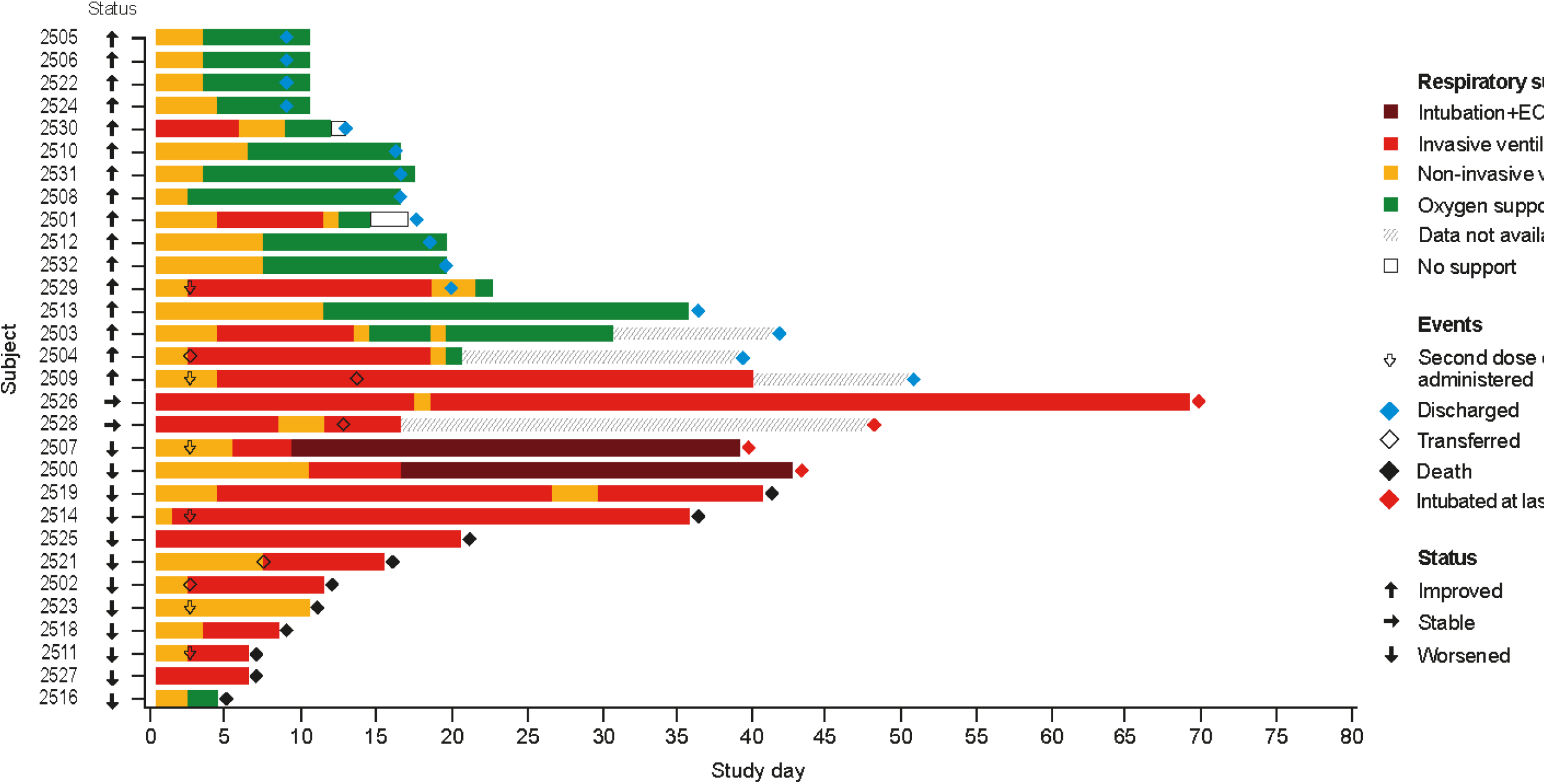
Changes in oxygen support and clinical outcome from day 1 through to study completion for all siltuximab-treated patients (n=30) ECMO=extracorporeal membrane oxygenation

### Laboratory parameters

Following treatment with siltuximab CRP levels reduced from a median of 20·7 mg/dL (IQR 12·85–25·30) at day 1 to a median of 0·3 mg/dL (IQR 0·1–3·9) at day 14 (figures 4a and 4b). Levels of liver function enzymes reduced over time in patients 30 days after treatment with siltuximab (AST: from 52·5 U/L at day 1 [IQR 40·5–74·5] to 24·5 U/L [IQR 16·5–40·0] at day 30). Lymphocyte, monocyte and leucocyte counts increased in patients following treatment with siltuximab (figure S2).

**Figure 4a:**
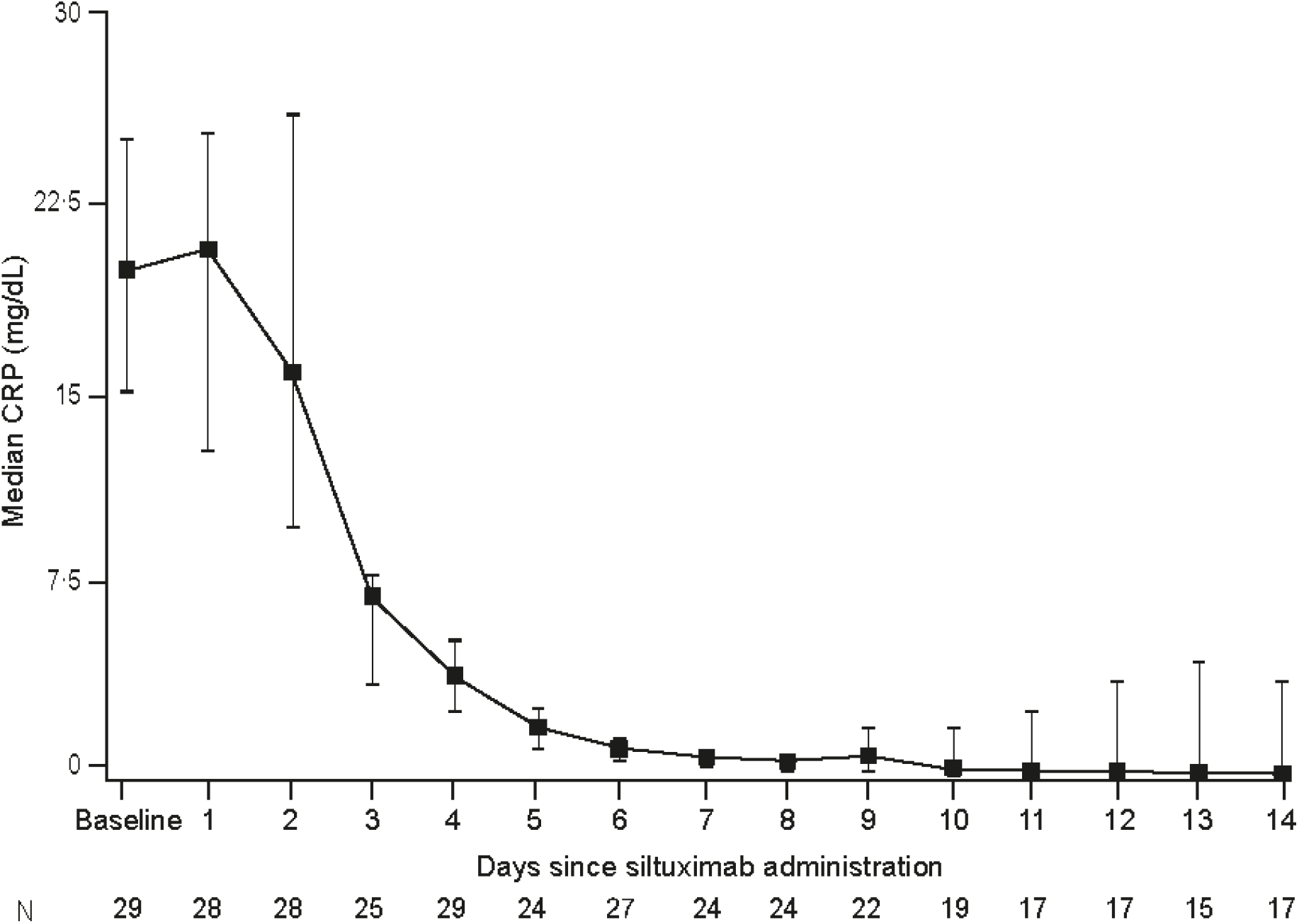
Median C-reactive protein levels from baseline to day 14 in all siltuximab-treated patients

**Figure 4b:**
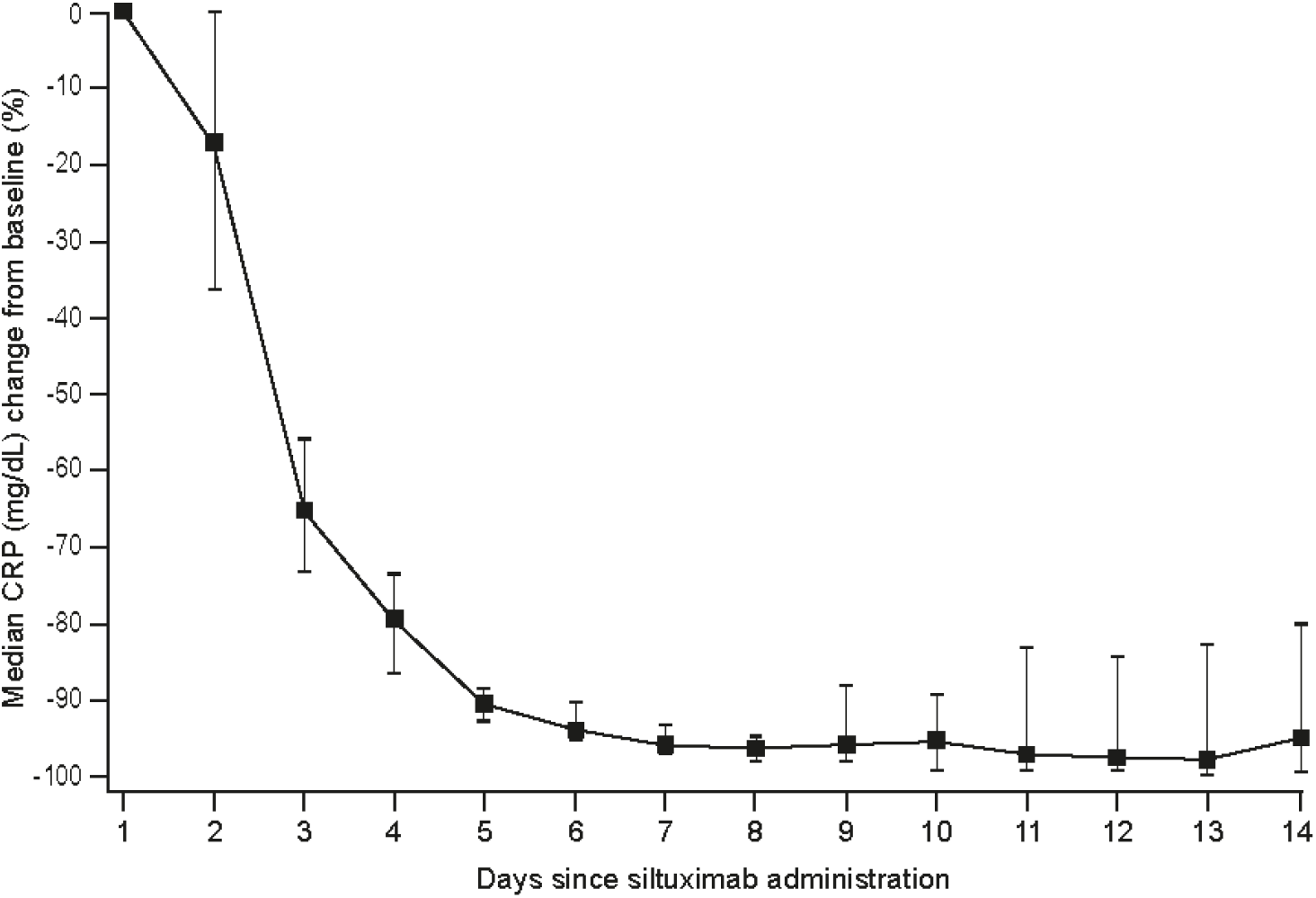
Percentage change in C-reactive protein from day 1 to day 14 in all siltuximab-treated patients

We completed a logistic regression analysis on IL-6 and although not significant, there was a trend developing between IL-6 levels and mortality (figure S3).

### Adverse events

No new or unexpected drug-related adverse events were reported in siltuximab treated patients, and the majority were Grade 3 or less. Table 3 summarises the adverse events, and table 4 summarises the adverse event gradings. There were 10 deaths among siltuximab-treated patients: eight deaths were due to respiratory failure, one to multi-organ failure, and one to septic shock. The patient who died of septic shock was transferred to another hospital during the follow-up period. Of the 30 patients in the matched-control cohort, 16 patients died (cause of death data were not available).

**Table 3:** Summary of adverse events in siltuximab-treated patients.

|  | **Siltuximab-treated patients, IMV**<br>**n=5** | **Siltuximab-treated patients, CPAP/NIV**<br>**n=25** | **All siltuximab patients**<br>**n=30** |
| --- | --- | --- | --- |
| **At least one TEAE** | 5 (100·0%) | 21 (84·0%) | 26 (86·7%) |
| **At least one SAE** | 2 (40·0%) | 9 (36·0%) | 11 (36·7%) |
| **At least one severe TEAE** | 5 (100·0%) | 21 (84·0%) | 26 (86·7%) |
| **Deaths** | 2 (40·0%) | 8 (32·0%) | 10 (33·3%) |
| **At least one AE of CTCAE grading Mild** | 0 (0·0%) | 3 (12·0%) | 3 (10·0%) |
| **At least one AE of CTCAE grading Moderate** | 2 (40·0%) | 11 (44·0%) | 13 (43·3%) |
| **At least one AE of CTCAE grading Severe** | 4 (80·0%) | 20 (80·0%) | 24 (80·0%) |
| **At least one AE of CTCAE grading Life Threatening** | 2 (40·0%) | 4 (16·0%) | 6 (20·0%) |
| **At least one AE of CTCAE grading Death** | 2 (40·0%) | 8 (32·0%) | 10 (33·3%) |

AE=adverse event. CTCAE=Common Terminology Criteria for Adverse Events. SAE=serious adverse event. TEAE=treatment-emergent adverse event.

**Table 4:** Summary of adverse events of special interest stratified by grade (siltuximab-treated patients only)

| System organ class | Preferred term | CTCAE Grade |  |  |  |  |
| --- | --- | --- | --- | --- | --- | --- |
|  |  | 1 | 2 | 3 | 4 | 5 |
| **Patients with at least one event** |  | 2 (6.7%) | 13 (43.3%) | 24 (80.0%) | 6 (20.0%) | 10 (33.3%) |
| **Investigations** |  | **0 (0)** | **0** | **13 (43.3%)** | **0** | **0** |
|  | Alanine aminotransferase increased | 0 | 0 | 12 (40%) | 0 | 0 |
|  | Aspartate aminotransferase increased | 0 | 0 | 3 (10%) | 0 | 0 |
|  | Lipase increased | 0 | 0 | 1 (3.3%) | 0 | 0 |
|  | Platelet count decreased | 0 | 0 | 1 (3.3%) | 0 | 0 |
| **Infections and infestations** |  | **0** | **2 (6.7%)** | **8 (26.7%)** | **2 (6.7%)** | **1 (3.3%)** |
|  | Bacterial sepsis | 0 | 0 | 4 (13.3%) | 0 | 0 |
|  | Pneumonia, bacterial | 0 | 0 | 2 (6.7%) | 0 | 0 |
|  | Urinary tract infection | 0 | 2 (6.7%) | 0 | 0 | 0 |
|  | Bronchopulmonary aspergillosis | 0 | 0 | 1 (3.3%) | 0 | 0 |
|  | Encephalitis, viral | 0 | 0 | 1 (3.3%) | 0 | 0 |
|  | Pneumonia | 0 | 0 | 1 (3.3%) | 1 (3.3%) | 0 |
|  | Pneumonia, *Pseudomonas* | 0 | 0 | 2 (6.7%) | 0 | 0 |
|  | Septic shock | 0 | 0 | 0 | 1 (3.3%) | 1 (3.3%) |
| **Vascular disorders** |  | **0** | **2 (6.7%)** | **7 (23.3%)** | **1 (3.3%)** | **0** |
|  | Hypertension | 0 | 1 (3.3%) | 5 (16.7%) | 0 | 0 |
|  | Hypotension | 0 | 0 | 2 (6.7%) | 1 (3.3%) | 0 |
|  | Peripheral artery haematoma | 0 | 1 (3.3%) | 0 | 0 | 0 |
| **Respiratory, thoracic, and mediastinal disorders** |  | **2 (6.7%)** | **2 (6.7%)** | **6 (20.0%)** | **1 (3.3%)** | **8 (26.7%)** |
|  | Respiratory failure | 0 | 0 | 0 | 0 | 8 (26.7%) |
|  | Pulmonary embolism | 0 | 1 (3.3%) | 3 (10.0%) | 1 (3.3%) | 0 |
|  | Pneumomediastinum | 1 (3.3%) | 0 | 0 | 0 | 0 |
|  | Pneumothorax | 1 (3.3%) | 1 (3.3%) | 2 (6.7%) | 0 | 0 |
|  | Pulmonary haemorrhage | 0 | 0 | 1 | 0 | 0 |
| **Psychiatric disorders** |  | **0** | **5 (16.7%)** | **3 (10.0%)** | **0** | **0** |
|  | Delirium | 0 | 4 (13.3%) | 3 (10.0%) | 0 | 0 |
|  | Confusional state | 0 | 1 (3.3%) | 0 | 0 | 0 |
| **Renal and urinary disorders** |  | **0** | **0** | **5 (16.7%)** | **1 (3.3%)** | **0** |
|  | Acute kidney injury | 0 | 0 | 4 (13.3%) | 1 (3.3%) | 0 |
|  | Haematuria | 0 | 0 | 1 (3.3%) | 0 | 0 |
| **Gastrointestinal disorders** |  | **0** | **0** | **3** | **0** | **0** |
|  | Small intestinal haemorrhage | 0 | 0 | 2 (6.7%) | 0 | 0 |
|  | Gastric haemorrhage | 0 | 0 | 1 (3.3%) | 0 | 0 |
| **Skin and subcutaneous tissue disorders** |  | **0** | **2 (6.7%)** | **0** | **0** | **0** |
|  | Rash, maculo-papular | 0 | 2 (6.7%) | 0 | 0 | 0 |
| **Blood and lymphatic system disorders** |  | **0** | **0** | **0** | **1 (3.3%)** | **0** |
|  | Disseminated intravascular coagulation | 0 | 0 | 0 | 1 (3.3%) | 0 |
| **Cardiac disorders** |  | **0** | **1 (3.3%)** | **0** | **1 (3.3%)** | **0** |
|  | Atrial fibrillation | 0 | 1 (3.3%) | 0 | 0 | 0 |
|  | Bradycardia | 0 | 0 | 0 | 1 (3.3%) | 0 |
| **General disorders and administration site conditions** |  | **0** | **1 (3.3%)** | **0** | **0** | **1 (3.3%)** |
|  | Hypothermia | 0 | 1 (3.3%) | 0 | 0 | 0 |
|  | Multiple organ dysfunction syndrome | 0 | 0 | 0 | 0 | 1 (3.3%) |
| **Nervous system disorders** |  | **0** | **1 (3.3%)** | **2 (6.7%)** | **0** | **0** |
|  | Peripheral motor neuropathy | 0 | 0 | 1 (3.3%) | 0 | 0 |
|  | Transient ischaemic attack | 0 | 1 (3.3%) | 0 | 0 | 0 |
|  | Cerebrovascular accident | 0 | 0 | 1 (3.3%) | 0 | 0 |

CTCAE=Common Terminology Criteria for Adverse Events.

One serious adverse event, a Grade 3 cerebrovascular event, was reported in siltuximab-treated patients. A total of 13 siltuximab-treated patients reported an infection: two were Grade 4 and the remaining 11 were either Grade 2 or 3. Elevations that occurred in AST and ALT were all Grade 3 and resolved by the end of the study.

## Discussion

Our study is the first to report comparative data with control patients on the use of an IL-6 neutralising mAb, siltuximab, in the treatment of patients diagnosed with COVID-19 and respiratory failure. This observational cohort study included 30 patients treated with siltuximab, with 30 control patients who received best supportive care matched using a propensity score analysis to minimise differences in the baseline covariates.

At the time of designing this study, there was an urgent need for an effective treatment for patients with SARS-CoV-2 pneumonia, hyperinflammation, and respiratory failure.^16^ To date, many of the drugs employed to treat COVID-19 limit viral replication; however, since IL-6 drives the hyperinflammation associated with severe and critical COVID-19, inhibiting this inflammatory cytokine may control the pathological immune response.^17^ Results from observational studies of tocilizumab in patients with COVID-19 often requiring oxygen support rather than ventilatory support, have been mixed, with some studies showing an improvement in respiratory function, and others reporting no improvement in mortality.^11,12,18,19^ Although both tocilizumab and siltuximab target the IL-6 signalling pathway, their mechanism of action differs – tocilizumab is an IL-6 receptor blocker, whereas siltuximab directly neutralises IL-6, one of the key pro-inflammatory cytokines driving hyperinflammation associated with severe and critical COVID-19.^5,6,20^

Elevated levels of pro-inflammatory cytokines and disease markers, IL-1, IL-6, IL-8, lactate dehydrogenase, ferritin and CRP, alongside low levels of lymphocytes, have been shown to correlate with clinical severity in COVID-19.^5,6,21^ IL-6 and CRP were elevated in the siltuximab-treated patients at baseline in our study, indicative of hyperinflammation. Although hyperinflammation is a key driver of the tissue damage in COVID-19 it is still unclear when the use of immune blockers is beneficial, and whether if used too early their use could result in the development of secondary infections.^5,22^ In our study, patients with rapidly progressing, severe ARDS and elevated cytokines were treated with siltuximab when ventilatory support was required. Mortality rates can reach over 85% in patients requiring IMV, and therefore preventing progression of respiratory failure in patients with COVID-19 is an urgent clinical unmet need.^23^ Following treatment with siltuximab, the risk of death was reduced by 54% in patients on either CPAP/NIV or IMV compared with the matched-control cohort with up to 42 days of follow-up (HR=0·462; p=0·0399). There was a trend towards a reduction in the time to IMV or death in patients on CPAP/NIV treated in the siltuximab and matched-control cohorts, but the result was not significant. Previous studies have shown that IL-6 levels correlate with outcome; however, in our study although there was a trend towards a correlation between IL-6 levels and mortality in siltuximab-treated patients, the result was not significant, perhaps as all patients treated with siltuximab had severe disease with high IL-6 levels.^8,9^

In addition to improvements in mortality, laboratory data show a reduction in inflammation as evidenced by a rapid and sustained fall in serum CRP. The reduction in CRP suggests that siltuximab neutralised IL-6, as CRP is an acute-phase protein regulated by IL-6 and is an established surrogate marker.^24^ In addition, the normalisation of serum CRP from Day 5 onwards coincides with the observed separation of the survival curve, which supports findings that increased serum CRP and IL-6 are linked with mortality in COVID-19.^8,9^ Following treatment with siltuximab, more than 50% of patients were discharged from hospital, the majority within 25 days of the follow-up period, and four within 55 days of treatment. The number of patients discharged in our study are similar to those reported in the remdesivir compassionate-use study (remdesivir 47% and siltuximab 53%).^25^ In the Campochiaro et al. study, 63% of tocilizumab patients were discharged within 28 days of the study.^26^

Siltuximab was well tolerated and the adverse event profile in patients with COVID-19 was similar to that reported in patients with idiopathic multicentric Castleman disease.^27^ No adverse events were considered to be related to the study drug. Elevations in the liver function enzymes AST and ALT are often present at baseline in patients with COVID-19, and in siltuximab-treated patients all elevations were Grade 3 and resolved to normal levels over time.^28^ Rates of secondary infections were apparently lower than those reported in some studies with tocilizumab (13–70% *vs* 43% in our study).^19,22,26^ In one study, tocilizumab was associated with a significant increase in secondary bacterial infections, including hospital-acquired pneumonia and ventilator-associated pneumonia.^22^ Those patients who did develop bacterial secondary infections in our study were mostly classed as having a Grade 3 infection, and resolved with one exception: one patient with bacteraemia progressed to septic shock and died during the follow-up period. Lymphocytes, leucocytes and monocytes counts increased following treatment suggesting that recovery of immune function was initiated, possibly restoring the host antiviral response.^5,29^

Two patients in each group received remdesivir (data not shown) as antiviral drugs were permitted as best supportive care; however, since remdesivir was given to so few patients and is most active in the early stages of COVID-19 it is unlikely to have impacted the treatment outcomes in this study.^30^

Our study has several limitations. It is an observational study with a small sample size, which limits the interpretation and generalisability of the results. Due to the observational study design, we cannot exclude the possibility of unmeasured confounding factors, although we have reassuringly noted consistency in the propensity score-matched analyses. No data were available for laboratory parameters and adverse events in the control cohort, meaning that comparisons between treated and untreated patients were not possible. Nonetheless, there is a sound rationale for the use of siltuximab in this patient group, and there were improvements in mortality and patient outcomes.

In summary, our study indicates that siltuximab administered at the onset of ventilatory support reduces mortality associated with COVID-19 and respiratory failure compared with best supportive care. Further randomised controlled studies are needed to confirm the efficacy and safety of this IL-6 neutralising mAb in the treatment of patients with COVID-19 and respiratory failure.

## Data Availability

All data will be made available upon reasonable requests submitted to the authors.

## Acknowledgments

We would like to thank the patients who volunteered to participate in this study. EUSA Pharma provided the study drug, and funded the data collection, analysis, and interpretation. Data collection and analysis was completed by Eva E Alder of ErgoMed. Medical writing assistance was provided by Cheryl Jenkins of TVF Communications (London, UK).

